# Content quality versus sharing practices on social media: A cross-sectional analysis of nutrition information on Twitter

**DOI:** 10.1101/2024.08.15.24312059

**Authors:** Cassandra H. Ellis, Peter Ho, J Bernadette Moore, Charlotte E.L. Evans

## Abstract

**Objective:** To use the validated Online Quality Assessment Tool (OQAT) to assess; the quality of online nutrition information, the difference between re-tweeted articles, and the impact of original information source.

**Setting:** Twitter (X) as little known about the quality of nutrition information shared on the platform.

**Design:** Tweet Archiver collected tweets including the word ‘nutrition’ on seven randomly selected days in 2021. Tweets were screened for URL inclusion and grouped based on retweet status. URLs were assessed using the OQAT. Rasch measures defined quality levels (low, satisfactory, and high-quality), while mean differences of retweeted and non-retweeted data were assessed by the Mann-Whitney U test. The Cochran-Mantel-Haenszel test was used to compare information quality by source.

**Results:** In total, 10,573 URLs were collected from 18,230 tweets. After screening for relevance, 1,005 articles were assessed (9,568 were out of scope) sourced from: professional-blogs (n=354), news-outlets (n=213), companies (n=166), personal-blogs (n=120), NGOs (n=60), magazines (n=55), universities (n=19), government (n=18). Rasch measures indicated the quality levels; 0-3.48, poor, 3.49-6.3, satisfactory and, 6.4-10, high quality. Personal and company-authored blogs were more likely to rank as poor quality. There was a significant difference in quality of retweeted (n=267, sum of rank, 461.6) and non-retweeted articles (n=738, sum of rank, 518.0), U = 87475, p=0.006, but no significant effect of information source on quality.

**Conclusions:** **Lower-**quality nutrition articles were more likely to be retweeted. Caution is required when using or sharing articles, particularly from companies and personal blogs, which tended to be lower-quality sources of nutritional information.

## Introduction

It is becoming increasingly more common for the public to turn to the internet and social media sources for nutrition information ^(1)^. However, the digital environment has minimum regulation and the quality of information can vary greatly ^(2)^, which puts the public at risk of exposure to conflicting nutrition information or misinformation online ^(3)^. Any individual, or company, can create and share content online, regardless of their expertise, with lack of quality control increasing the risk of knowledge distortion ^(4)^. To add to the complexity, social media facilitates rapid dissemination of content ^(5)^ allowing myths to spread quickly ^(6)^ and creating an environment where ‘often the loudest, most extreme voices drown out the well informed’ ^(7)^.

Previous research has assessed the quality of different types of nutrition information online, but the number of studies is small, the quality assessment criteria variable, and search terms often relate to specific dietary choices. For example, one study found variation in the quality of online articles giving information on vegan diets, with 45% of articles rated poor or very poor, compared to only 9% being rated as excellent ^(2)^. Others have assessed the quality of websites providing consumer information on dietary and herbal supplements for weight loss, finding that the highest scoring websites were dedicated health portals, and the lowest were commercial websites ^(8)^.

In recent years there has been a proliferation of professional bloggers giving lifestyle and dietary advice ^(9; 10)^. In the context of nutrition, many bloggers have thousands of followers, but no nutritional science training or relevant qualifications ^(6)^. Indeed, it has been found that only 6% of American food bloggers have nutrition degrees, despite offering nutrition advice ^(11)^. Analysis of popular Australian healthy eating blogs found that blogs written by professionals were higher quality, with 64% giving healthy eating information in a practical manner. However, only 43% of the blogs aligned with advice from the Australian Dietary Guidelines ^(10)^. Supporting this, personal blogs have been found to consistently be of poorer quality than other sources of online information ^(2; 12)^, and commercial articles providing adolescents with lifestyle and nutrition advice found to be subjective and unbalanced ^(13)^. In part, this could be explained by UK Article 12(c) in the Nutrition and Health Claims Regulation ^(14)^, which prohibits health professionals discussing certified health claims in commercial communications. However non-professionals, celebrities, and ‘influencers’ do not fall within this regulation and can discuss health claims, whether certified or not ^(15)^.

In recent years, online articles giving advice on COVID-19 and vitamin D were found to have provided conflicting information, inconsistent with the scientific evidence ^(16)^. In addition, articles giving clinical nutrition advice to patients were shown to vary greatly in quality. For example, websites providing nutrition advice to anorexia nervosa patients were determined to be too complex, with important information on the benefits and risks of treatment options not covered ^(17)^. Moreover, similar patterns of poor-quality nutrition information being disseminated by non-experts has been evidenced on social media, with 94% of Instagram posts giving nutrition advice assessed as ‘extremely low’ quality and less than 1% as high quality ^(18)^. A separate study also using Instagram, found weight management blogs by social media influencers to be of poor quality, with most scoring less than 50% against defined quality criteria ^(19)^. Whereas, YouTube videos giving dietary advice after bariatric surgery were found to be of higher quality when uploaded by physicians and dieticians, but lower quality videos by non-experts had more views and likes ^(20)^. Relatedly, the ‘healthy diet’ discourse on Twitter has been found to be dominated by ‘non-health professionals’ and largely constitutes poor quality information contradictory to public health advice ^(5)^. This is particularly concerning as 77% of adolescents in an Australian study reported using social media for healthy lifestyle information ^(13)^.

However, it is difficult to compare the quality across these existing studies due to the use of multiple quality criteria. Moreover, Afful-Dadzie and colleagues conducted a literature review on the quality of health information shared on social media and found that most of the literature relied on three tools that were created before the widespread use of social media ^(21)^. Afful-Dadzie et al concluded these tools were outdated and not fit for purpose, leading them to call for standardised quality assessment suitable for social media and online content. This led the authors of this paper to develop and validate a tool, specifically suited to assessing the quality of online nutrition information, which will eventually allow comparison between studies using the tool.

Examining social media can also provide unique insights into the nutrition and diet information reaching, and influencing, large segments of the general population ^(22)^. In addition, sharing practices are important to understand as Twitter allows for the initial sharing of a post, but this post is also subject to secondary shares, or retweets, and it has been found that three in ten people who share links to articles on Facebook or Twitter, do so without reading the content first ^(23)^. Previous research has investigated emotion as a motivator for retweeting news ^(24)^, but to our knowledge, the quality of the information being shared, and whether high quality information is more likely to be retweeted, has not been investigated. Poor quality online communication is prevalent in other disciplines; it can amplify political misinformation^(25)^, encouraging unconstructive discussion^(26)^ and causing government authored content to gain little attention ^(27)^. Public authored blogs on climate change are less likely to be evidence based^(28)^ and can have a negative impact on the public debate. Therefore, in the context of widespread sharing of misinformation, it is important to understand the quality of the information that has the potential to be widely shared^(29)^.

In summary, nutrition research is at particular risk of misunderstanding as people have daily interactions with food, and beliefs may be rooted in cultural practices, assumption and intuition, more than sound science^(30)^. Prolonged exposure to inconsistent nutrition information over a period of time can have detrimental effects on consumer beliefs^(31)^,^(7)^ and impact adherence to recommended nutrition behaviours such as fruit and vegetable consumption^(32)^. Therefore, it is increasingly important to be able to differentiate between high- and low-quality nutrition information and determine the sharing practices of different types of information.

This study aimed to address a gap in the literature by using a newly developed and validated online quality assessment tool ^(33)^ to assess the quality of online nutrition information disseminated via Twitter. It aimed to examine the quality of retweeted articles, comparing these to unshared content and determine whether the high- or low-quality information is more likely to be retweeted, and determine which sources were sharing the highest quality nutrition information.

## Methods

Using a previously validated tool designed to measure the quality of online nutrition information^(12)^ ^(33)^, we aimed to analyse the quality of a randomly selected subset of nutrition related articles posted on Twitter in 2021. Whilst Twitter changed its name to X in July 2023, the data collected for this study was collected from Twitter, therefore we will continue to refer to the platform as Twitter and use the terms tweets and retweets throughout this study.

### Data collection and screening

Tweet Archiver ^(34)^ was used to automatically webscrape a dataset of all English language tweets that included the word ‘nutrition’ by month from 1 January 2021 to 31 December 2021. A full year was collected to allow a random sample from across the year to be analysed which would not be affected by any predetermined seasonal effects, usually seen in December and January^(35)^.

Using Random.org, four days were selected for analysis, 24 January, 11 August, 21 November and 22 November 2021. There were more tweets collected that had not been retweeted, therefore three additional days were randomly selected: 26 May, 12 June and 14 December 2021, and the retweeted tweets were included for analysis. This gave approximately the same number of URLs in each category (retweet and no retweet) before screening for relevance.

The data were then filtered by those containing a URL, tweets that did not include a URL were discounted. This established two datasets: URLs with and without retweets.

Each eligible URL was reviewed manually and categorised based on the OQAT codebook^(12)^ to identify the website source and the content type. URLs were excluded if topically irrelevant, linked to social media or consisted of advertising and product promotion. Articles that related to climate change, animal nutrition, food and agricultural policy were discounted if they did not directly relate to nutrition and human health. In addition, URLs were discounted if they were part of discussion forums, videos, or linked to other social media accounts as the OQAT was only designed to measure written information.

Two trained raters used the tool independently to score the relevant URLs against the 10 OQAT criteria. A higher OQAT score indicated a higher quality article. Any discrepancies were discussed and consensus was reached. After scoring, articles were ranked into three categories using the OQAT measure obtained from the Rasch analysis described in the next section. The source of the URL was also recorded by the OQAT. Articles were manually categorised by raters and categorised as one of the following 10 sources; 1 Blog – personal, 2 Blog – professional, 3 Company, 4 Government organisation, 5 Magazine, 6 Non-Governmental Organisation (NGO), 7 Professional news, 8 Research institute/University/publisher, 9 social media (out of scope), 10 unrelated (out of scope). Raters met to discuss and agree any ambiguity. Rater one checked a random sample of rater two’s scores to ensure correct application of the OQAT, any discrepancies were discussed and agreed. Rater reliability was checked using Rasch model; results are presented in the supplementary material.

### Statistical analysis

The Statistical Package for the Social Sciences (SPSS v 28.0) was used for statistical analysis and the R computing environment (v 4.2.3) was used for data visualisation. After all tweets including the word ‘nutrition’ posted in 2021 were collected, tweets were collated and those including a URL were identified. The raw data was charted to visualise the annual data collection. The data collect and screening was visualised in a flowchart. Descriptive tests were reported including total scores, medians and Interquartile Ranges were calculated for each media source and by retweet.

A total measure for evaluating quality was obtained by fitting a Rasch rating scale model to the 10 item OQAT questionnaire. Quality levels (low, satisfactory, high) were established by determining statistically significant levels in the Rasch measures based on the procedures suggested by Wright ^(36)^. Prior to determining the quality levels, data evaluated by each rater was also examined with a separate Rasch model, to confirm that both raters were using the OQAT tool in a similar manner, so that data could be combined in a single analysis. Standard procedures for checking item fit and other Rasch model assumptions were conducted as described^(37)^.

As the data was categorical, the Cochran-Mantel-Haenszel test ^(38)^ was used to examine the associations between high, satisfactory and low-quality articles, and whether they were more of less likely to be retweeted. The contingency analysis is displayed as a Fourfold graph to allow the categorical data to be visualised. The Woolf test was used to test the homogeneity between log odds ratios in each strata to determine whether the Cochran-Mantel-Haenszel test was valid. Cochran-Mantel-Haenszel test was also used to investigate whether there was a significant difference between sources, when comparing by whether they were retweeted. Cochran-Mantel-Haenszel test was chosen as it is more robust when some of the strata contain small frequencies. After the contingency and chi-square analysis, URLs were manually reviewed by rater one to see whether it was possible to infer any rationale for differences between groups.

The Shapiro-Wilks Test was used for normality of retweets and non-retweeted data indicated that the data were not normally distributed (p <0.001). The natural logarithm was used to transform the data but did not rectify the distribution and therefore non-parametric tests were used to compare tweets and retweets. The Mann-Whitney U test was used to analyse any differences in rank scores of retweeted and unshared data.

## Results

### Data collection

Over the full 12-month collection period, 943,869 tweets that included the word ‘nutrition’ were collected, and of these 591,907 contained an URL (figure 1). For quality analysis, four days were randomly selected for analysis, with an additional three days randomly selected for retweeted data analysis to ensure equal group sizes before analysis.

**Figure 1.**
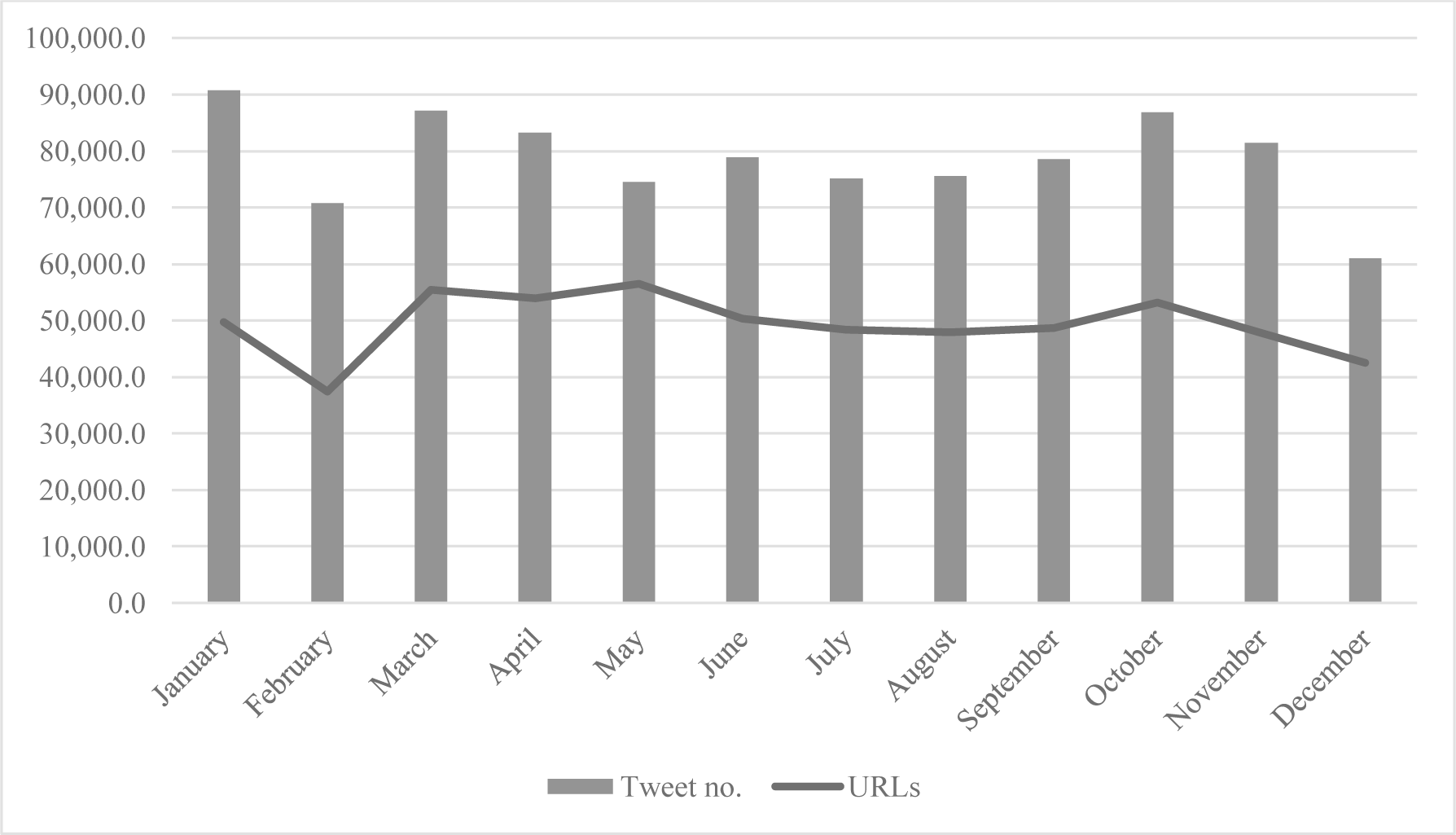
Total number of tweets categorised as nutrition information and URLs collected by month in 2021.

Over the 7-day analysis period, 10,573 URLs were collected from 18,230 Twitter posts. After manual screening for relevance to human nutrition and health, these represented: professional blogs n=354 (35.2%), news outlets n=213 (21.2%), companies n=166 (16.5%), personal blogs n=120 (11.9%), NGOs n=60 (6.0%), magazines n=55 (5.5%), research institutes or publishers n=19 (1.9%), government organisations n=18 (1.8%). URLs advertising, recipes, scientific papers, social media accounts, discussion forums or videos, 9,568 URLs were excluded as they were out of scope (figure 2).

**Figure 2.**
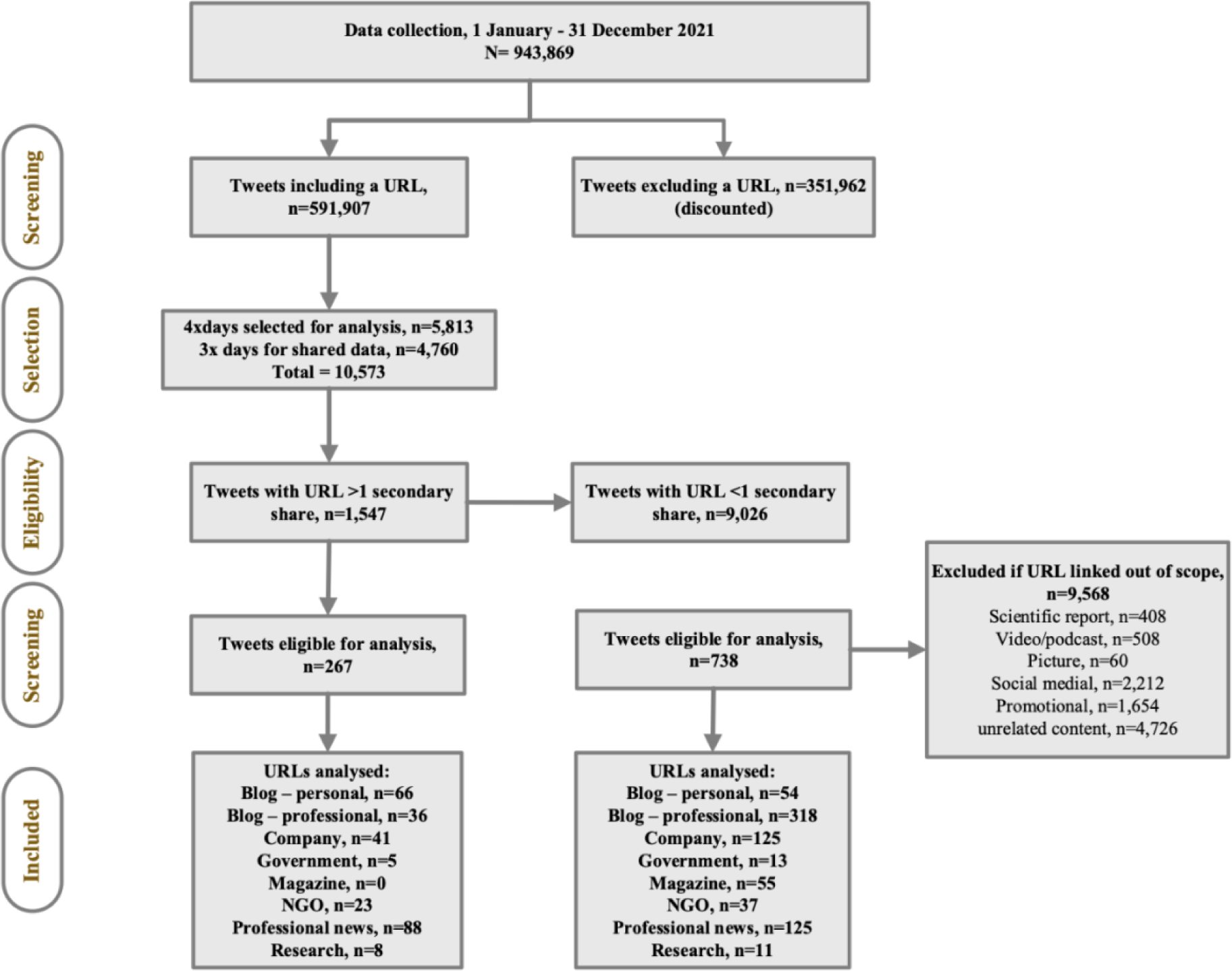
Flow diagram of identification and screening of tweets for analysis to assess the quality of online nutrition information.

### Fit and inter-rater analysis

Rasch analysis was conducted to ensure the OQAT criteria measured what they were designed to measure and to check inter-rater reliability. The Rasch analysis of the data indicated that all 10 items complied to the recommended OUTFIT mean squares between 0.5 – 1.5 for being “productive for measurement’^(36)^. Rasch analysis also confirmed that all sources met criteria 9, and criteria 4, 5 and 6 were necessary for an article to be classified as high. Figure three shows the fit with outliers removed for Q6 and Q9 to improve fit. Removing the outliers improved the fit but did not change the conclusions.

The Wright map (figure 3) shows the criteria (Q1 -Q10) ranked by prevalence, left to right. Details of what these criteria are designed to measure can be found in the authors previous paper ^(12)^. The lower plot indicates the criteria (Q1-10) by order of prevalence, left to right. Criteria on the left were more likely to be scored positively in the articles than those on the right. Therefore, we can see that all articles scored positively on Q9, and the least likely criterion to be met was Q6. The shading on the lower plot indicates the quality rank (low, satisfactory and high quality), therefore we are able to determine that; Q9, Q8 and Q10 were necessary for an article to score 3 and be deemed low quality, Q1, Q2, Q7 and Q3, were necessary for articles to be classified as satisfactory, and Q5 and Q6 were required for an article to be high quality.

**Figure 3.**
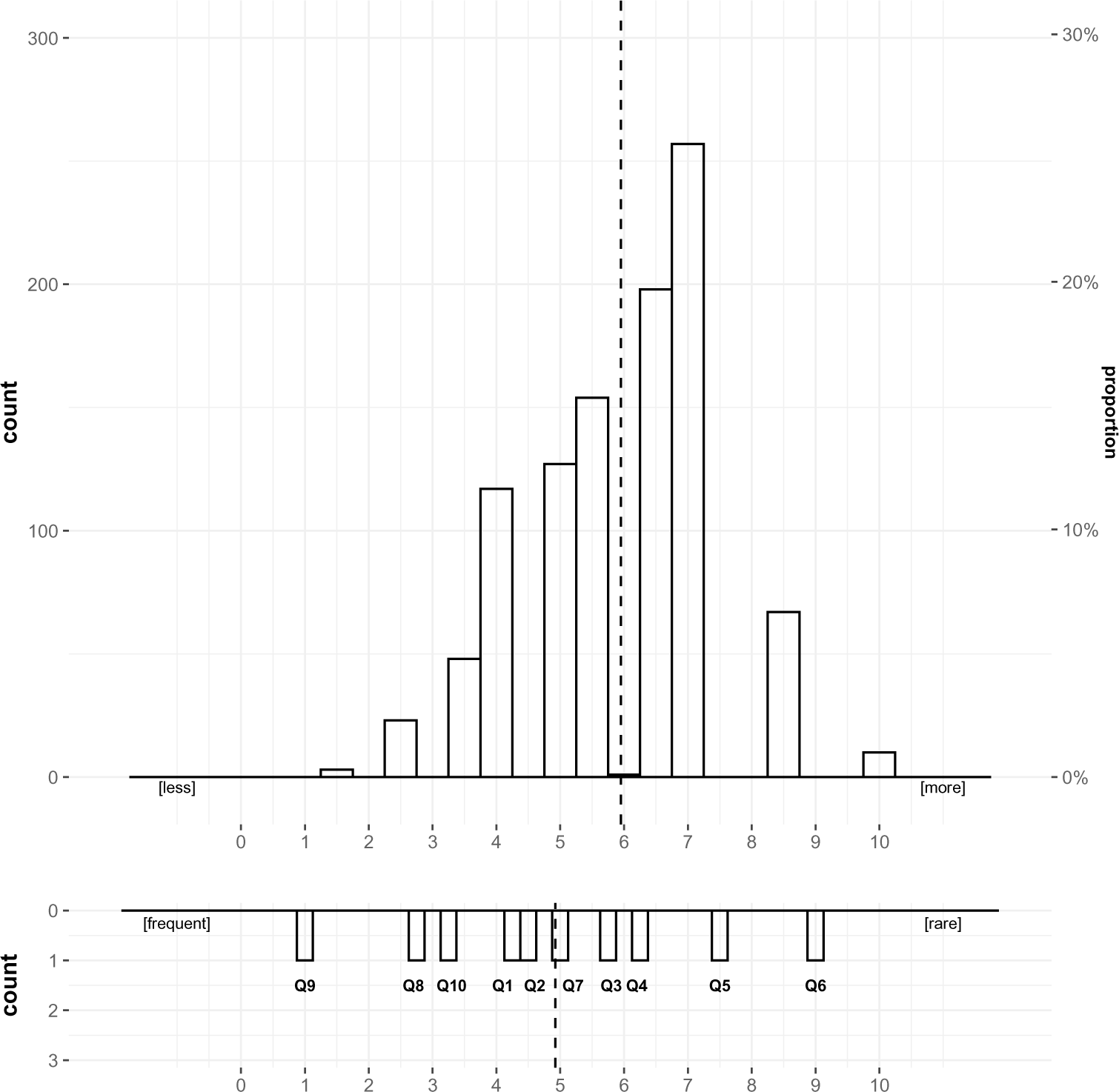
Wright map illustrating the quality of each article and discriminating quality assessment criteria. The upper plot shows the quality of each article. The lower plot illustrates the fit of all quality assessment criteria. Shaded areas from left to right of the plot correspond increasing levels of quality (low, satisfactory, high). All estimates were rescaled from 1-10. The dotted line represents the mean score.

To ensure inter-rater consistency, Rasch model was used to compare the two independent sets of rater scores. The distribution confirms that the value added to each criterion by each rater is the same inferring consistency between rater (supplement one).

### Descriptive analysis

To assess quality, articles were categorised as poor, satisfactory, and high quality based on the OQAT measure; 0-3.48 indicated poor quality, 3.49-6.3 indicated satisfactory quality and 6.4-10 indicated high quality. The quality levels are as identified by the OQAT using Rasch analysis which identified the minimum requirements for each category^(12)^.

The relevant URLs (n=1005) were assessed using the OQAT. As per the OQAT guidelines, 33% (n=335) of articles were categorised as high quality, 59% (n=595) as satisfactory, and 7% (n=74) were defined as poor quality articles (table 1).

**Table one.**
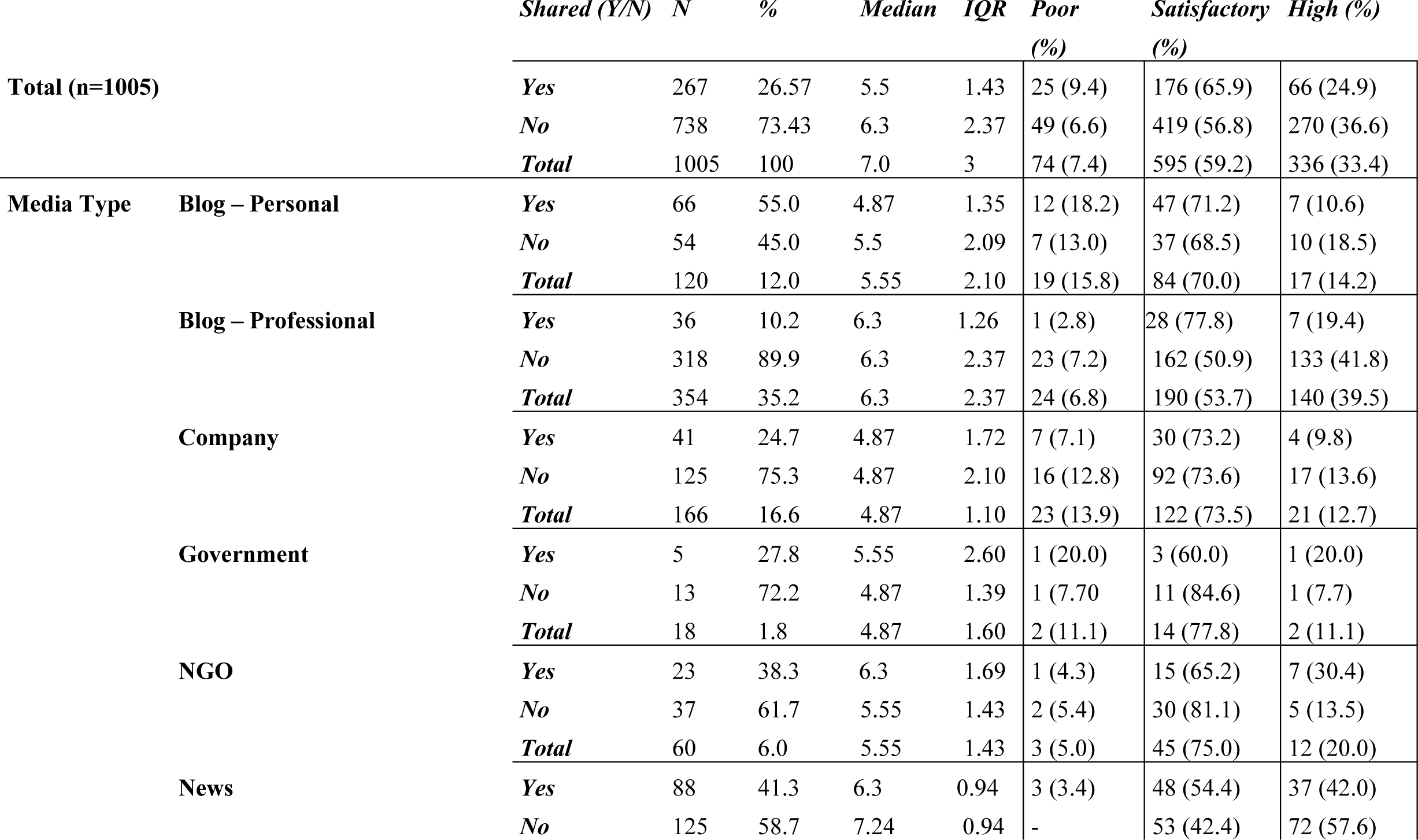
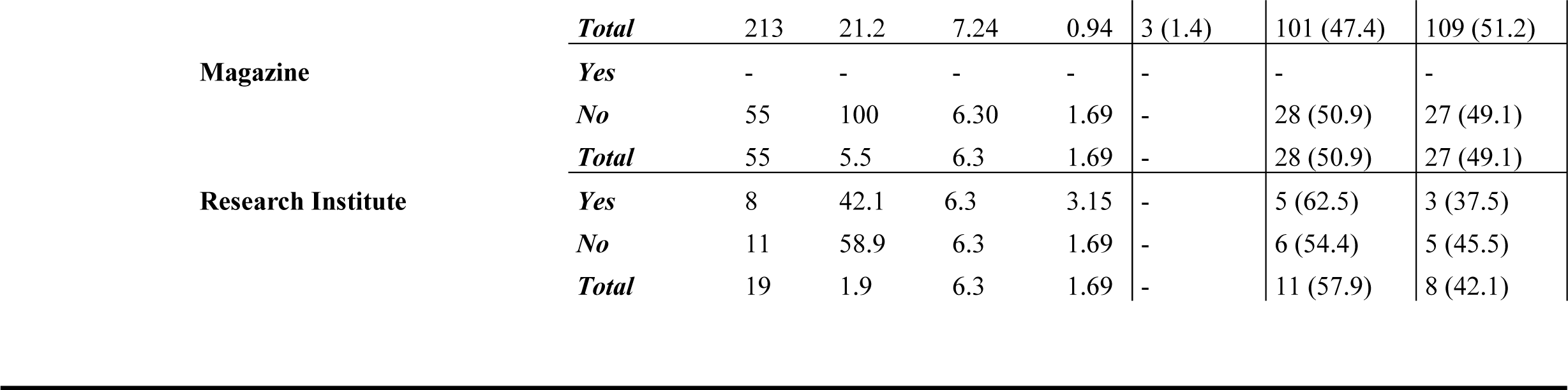
Online Quality Assessment Tool ranks by shared status, content type and media source.

### Retweet and no retweet comparison

URLs that were not retweeted (n=738, mean=6.03) scored higher on the OQAT than those that had been retweeted (n=267, mean=5.731). There was a significant difference in the quality of retweeted (n=267, sum of rank, 461.62) and non-shared data (n=738, sum of rank, 517.97), U = 87475, p=0.006. Articles categorised as poor and satisfactory by the OQAT, with a score of <6.3, were more likely to be retweeted. Similarly, articles defined as high quality had fewer retweets.

### Media source

The media source of the URL was recorded by the OQAT. The Woolf Test was used to test homogeneity of the logs ratio for each strata to ensure the Cochran-Mantel-Haenszel test assumptions were met and it was the most appropriate test, p=0.853. The mean scores for each media source were calculated with professional news outlets having the highest score, mean=6.67, and company blogs the lowest, mean=5.11 (table 1). When comparing retweeted and unshared by source, news had the highest mean score (retweeted 6.47, unshared 6.42), however, personal blogs had the lowest retweeted mean (4.92) and company blogs not retweeted has the lowest mean (5.14).

### Quality by media source

The Cochran-Mantel-Haenszel test was used to investigate whether there was a significant difference between sources, when comparing by whether they were retweeted. Results comparing high and satisfactory articles are displayed in figure four. When analysing the source quality, a comparison of high- and low-quality articles was also carried out (not presented) but because the group sizes of the low-quality articles was small, there was no significant difference, *X*^2^MH = 1.2487, df = 1, p = 0.264. Similarly, there was no significant difference between; satisfactory and low, *X*^2^ = 0.017, df = 1, p = 0.898; or between high and satisfactory, *X*^2^ = 0.888, df = 1, p = 0.346 (figure 4). The low numbers of retweets in most groups may have influenced the significance. Figure four shows the differences within groups, for example the government and research categories had low numbers of retweeted articles.

**Figure 4.**
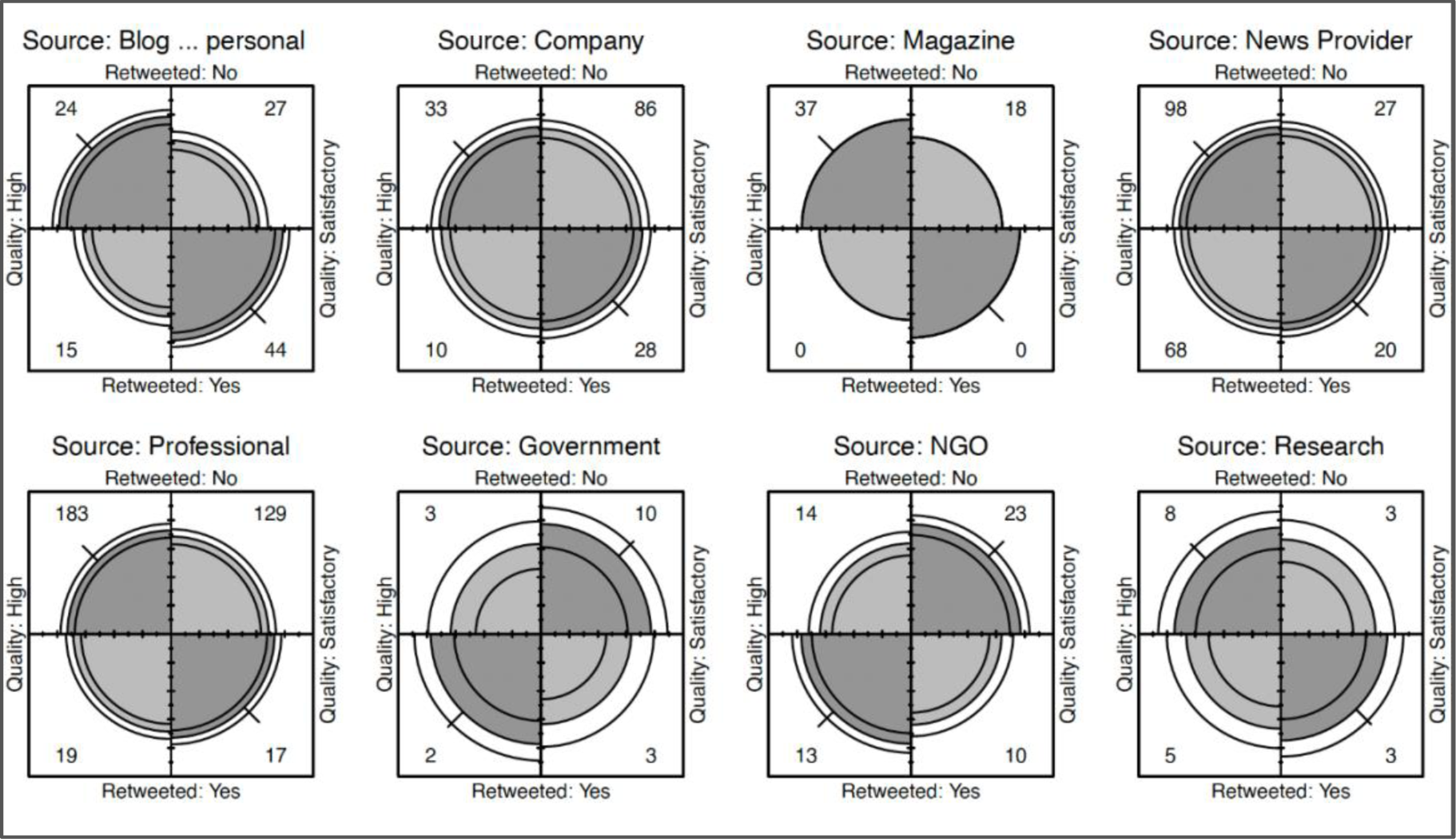
Fourfold display of article quality (High vs Satisfactory) by source. In each panel the darker shaded diagonal areas with greater area than the off-diagonal areas, show a positive association. The confidence rings for adjacent quadrants overlap if the odds ratio for quality and retweet does not differ significantly from 1.

## Discussion

In this study we measured the quality of a representative subset of public facing online nutrition information using a validated tool designed specifically for nutrition research; addressing an important gap in the literature. Importantly, we investigated whether Twitter users were more likely to retweet high- or poor-quality information and which media sources were more likely to share higher quality nutrition information. Our results show, for the first time, a significant difference in the quality of retweeted and non-retweeted nutrition articles, with lower quality content more likely to be retweeted.

There remains a paucity in the nutrition literature on whether the quality of information is a predictor of sharing, although poorer quality videos have been found to have more views and likes ^(20)^. Additionally, the lack of evidence based information retweeted in our study was consistent with the literature pertaining to anti-climate change blogs^(39)^ and public authored political blogs^(26)^. In this study, higher quality nutrition sources were less likely to be retweeted. Indeed, articles defined as poor or satisfactory were more likely to be retweeted. This suggests that either quality is not an important consideration for Twitter users when choosing to retweet, or that people are generally unable to discriminate between high- and low-quality nutrition information. As articles ranked satisfactory were the most retweeted, further investigation was carried out into whether users were more likely to retweet articles scoring high or low within the satisfactory range. There was not enough evidence to determine whether quality was a factor affecting retweet decisions for these users.

Analysis of climate change content shared on Twitter found that the accuracy of content does not impact sharing, rather novel content was more likely to be shared and retweeted ^(40)^. There is also another possibility, in that people do not read articles before sharing and therefore are not able to make an informed decisions on quality^(23)^. However, in this study more satisfactory than low quality articles were retweeted in this study suggesting that some quick ‘sense checks’ of quality may be taken before sharing. This aligns with previous research which suggests that some members of the public do engage in rapid checks to validate online health information before sharing ^(41)^.

Article quality varied greatly however poorer quality information was more likely to be retweeted than high quality. This supports previous work whereby online blogs scored poorly when measured against dietary advice. In particular, content which scored poorly was less likely to provide references to scientific evidence, provide expert quotes or to declare any author conflicts of interest ^(2; 10; 19)^. When comparing the quality of articles by source, the group sizes were not equal so calculating effect size was not possible. There appears to be a relationship between the source and the quality of the article, with commercial websites scoring lower, and professional news outlets scoring higher. This is supported by the literature. YouTube videos are higher quality when produced by experts, ^(20)^, and lifestyle websites written by commercial companies lack objectivity and transparency ^(13)^. Similarly, commercial websites giving advice on dietary supplements are more likely to be poor quality than those authored by health experts ^(8)^.

Interestingly, our results show magazine articles were unique in that they did not have any retweets, regardless of the quality of the article. In-depth analysis of these articles suggests that magazine articles may be distinctive in that they target a specific cohort such as women, marathon runners or vegans. The language used for magazine articles was simple and they were targeted towards the public, however they were more likely to give healthy eating advice to a specific group with specific requirements which may not have been novel enough to retweet ^(40)^. Articles targeting women may be less likely to be retweeted as fewer women use Twitter compared to men ^(42)^. Magazine articles were also more likely to be subscription based with access limited therefore people may be less willing to retweet content that their networks are unable to access.

A further novel finding was that articles shared by government agencies were also less likely to be retweeted than other sources ^(27)^, particularly if they were giving public health advice. Articles that related to population health and diets which were written for public health professionals were more likely to be retweeted. This could be due to academics and professionals being encouraged to use Twitter as a medium to disseminate research and network with peers. This could also be because the retweets were from other organisations, and these resources are therefore being used in a professional capacity, however this level of network analysis was out of scope for this study.

Including scientific references (Q4), quoting a specialist (Q5), and disclosing conflicts of interest or financial interests (Q6) were necessary criteria for articles to be deemed high quality. As shown in the Wright Map (figure 3), these essential criteria (Q4, Q5, Q6) where the least likely criteria to be achieved. This is consistent with the published literature whereby seeking expert opinion, a sign that the writer was concerned with fact checking, was lacking in many articles ^(43)^. The lack of evidence based information shared was also consistent with the literature pertaining to print news^(44)^, obesity^(45)^, and dietary advice to cancer survivors^(46)^. All of which highlighted the damage poor quality non-expert written information can have on public health and adherence to dietary guidelines. More encouragingly, the vast majority of articles scored positively on naming an author, an assessment criteria which has previously been shown to positively affect article quality^(43; 44)^. In our dataset, this criterion was necessary for an article to be ranked as satisfactory.

At the point of data collection, only five articles had greater than five retweets. This was notable as most retweeted articles had just one or two retweets. The most retweeted post was a high-quality article originally published by the World Food Programme, and originally tweeted by António Guterres, Secretary General of the United Nations. The next highest retweeted post was a link to a satisfactory article posted by a high-profile Twitter user with 4 million followers. Both of these Twitter users have large networks suggesting that the user network could be more influential than the quality of the article, however, as networks were not investigated in this research, we do not have enough information to confirm the influence of Twitter networks.

### Strengths and limitations

The main strength of this study is that it used a validated set of standardised assessment criteria^(12)^, as called for in the literature ^(13; 21)^, to assess the quality of nutrition information available online and shared on Twitter. By using a tool developed specifically to assess the quality of nutrition-related online content, our findings build upon recent studies that have categorised the positive characteristics of Dietitian authored blogs^(10)^, and compared the quality of the blogs to those from lay authors^(47)^. A further strength is the high inter-rater reliability. In this study, the two raters applied the OQAT consistently when rating the independent set of sources. In addition, to the authors knowledge, this is one of the only studies to quantify the quality of nutrition information by the source publishing the content.

Our data collection was novel in that it used Twitter as the source of URLs to objectively select a cross section of online articles designed to disseminate nutrition information. Therefore, each article analysed was interacted with at least once through the initial tweet reducing the likelihood of collecting passive content which does not stimulate reader engagement^(48)^. Additionally, these articles have increased chances of being viewed by the public as they are in the public domain in at least two formats, on the website and on Twitter. The random selection of days for analysis was a strength as it reduced the risk that the discourse was affected by seasonal variation ^(35)^.

However, there are some limitations to this study. The disproportionately lower number of retweeted articles compared to non-retweeted made comparison between groups difficult. Nonetheless, a greater number of nutrition related URLs not being retweeted is in line with the authors previous research investigating obesity articles online ^(45)^ and pilot studies using the OQAT^(12)^. In addition, the differing numbers between the sources limited comparison between these groups. Future research could also categorise articles differently comparing the quality of the type of content shared and not just the source.

A further limitation of the study methodology was that the raters were not blind to the article source. This could have introduced rater bias and caused the rater to moderate the article score based on subjective opinion. However, the OQAT criteria were worded as clearly as possible to reduce the risk of this type of bias, and inter-rater reliability was analysed to check that the OQAT was being applied consistently. In addition, only webpages were considered, therefore the wider limitations of the general website function are not considered. Similarly, this study did not consider article readability, as these can be assessed by external software such as Flesch-Kincaid readability test. Finally, only English language tweets and articles were included in the data set, so these findings may not be generalisable to tweets in other languages or non-English speaking countries. Approximately 40% of all tweets are written in English therefore a large proportion of nutrition related content was not considered in this research, and worthy of further exploration.

Although meta-data was collected, we are not able to infer motivations for retweeting, or any information about social networks. This is a limitation and an area for further research using social network theory to investigate Twitter networks, what users are sharing and retweeting, and who are the users sharing nutrition information. Similarly, this study did not consider which device users were sharing from so cannot make any inferences on whether users are more likely to share content on phones versus laptops, nor did we consider the feasibility of sharing through Twitter buttons on websites. However, future research considering the dissemination of content through networks could consider these factors.

Importantly, this research investigating the quality of information has led to a number of recommendations. Online content remains a popular source of nutrition advice for the public ^(10)^ ^(19)^, but the quality is variable^(45; 49; 50)^. In particular, recommendations to authors of online nutrition content are suggested. This research showed that to be considered high quality content, any article providing dietary advice must be evidence-based and include hyperlinks to the evidence or provide references. Hyperlinks and references must directly cite the evidence, and not opinion-based articles self-promoting other content on the same website. As digital content easily allows for hyperlinking content and an increasing proportion of nutritional journals are open access, it is proposed it is best practice to include scientifically validated weblinks.

Online content has an infinite lifespan, therefore must include a published date, and a review date. This was an essential criterion for articles to be considered of satisfactory quality. It is a necessary addition to ensure the reader can make informed decisions on the relevance and quality of the evidence presented and whether it includes out of date research. Another criterion required to be considered high quality is to include endorsements from specialists and subject matter experts. Expert quotes act as a mark of quality informing the reader that this is a well-researched article that has been subject to informal peer review. Finally, any funding or conflicts of interest should be explicitly stated for an article to be deemed high quality. This informs the reader of any potential author or publication bias, and again allows the reader to make an informed decision on whether the article is trustworthy. Further recommendations are included in the OQAT development and validation paper^(12)^.

### Conclusions and future research

The quality assessment of online nutrition information using a validated tool designed specifically for this purpose adds to a body of literature assessing quality of information in the media and online. This study contributes to the understanding of which sources of information the public are likely to engage with and what factors may motivate them to engage with it. Further research should consider using the OQAT on a larger data set. The current dataset is limited to one social networking site, Twitter, which does not capture all social media users and represents only one platform for sharing health information. Future research is needed that compares different public sources of nutrition and diet information and different social media platforms.

In addition, future research should consider the broader influences on retweeting beyond quality, with consideration given to the influence of the person posting the original tweet, the reach of their social media network and the influence of the site where the article is originally published. Finally, it is important for future research to explore the wider nutrition discourse on Twitter and the flow of information through networks to understand motivations for sharing nutrition content and who are the key actors.

## Data Availability

All data produced in the present study are available upon reasonable request to the authors

## Disclosure statements Acknowledgements

None

## Financial Support

This research received no specific grant from any funding agency, commercial or not-for-profit sectors.

## Conflict of Interest

None

## Authorship

CHE researched literature and conceived the study. CELE was involved in study design and protocol development. PH contributed to the statistical and data analysis. CHE wrote the first draft of the manuscript. All authors reviewed and edited the manuscript and approved the final version of the manuscript.

## Ethical Standards Disclosure

This study did not require ethical approval as all data was open source.

## Notes

### Competing Interest Statement

The authors have declared no competing interest.

### Funding Statement

This study did not receive any funding

