## Supplemental Figure 1a and 1b for "Content quality versus sharing practices on social media: A cross-sectional analysis of nutrition information on Twitter"

**Table of Contents**

Figure 1a Wright Map of Rater One scores
Figure 1b Wright Map of Rater Two scores

### Supplement 1 Scores by rater

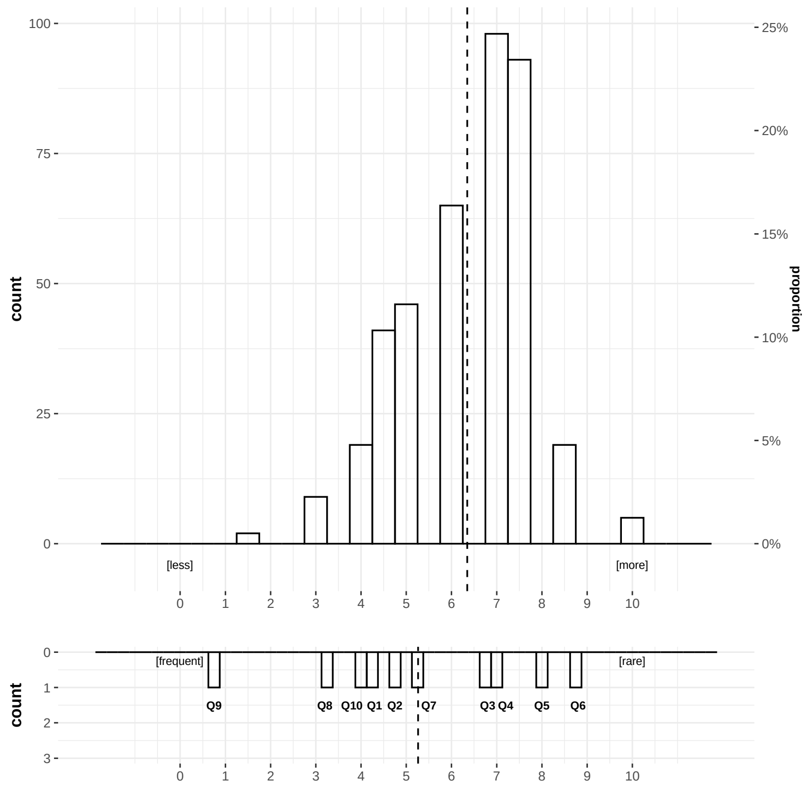


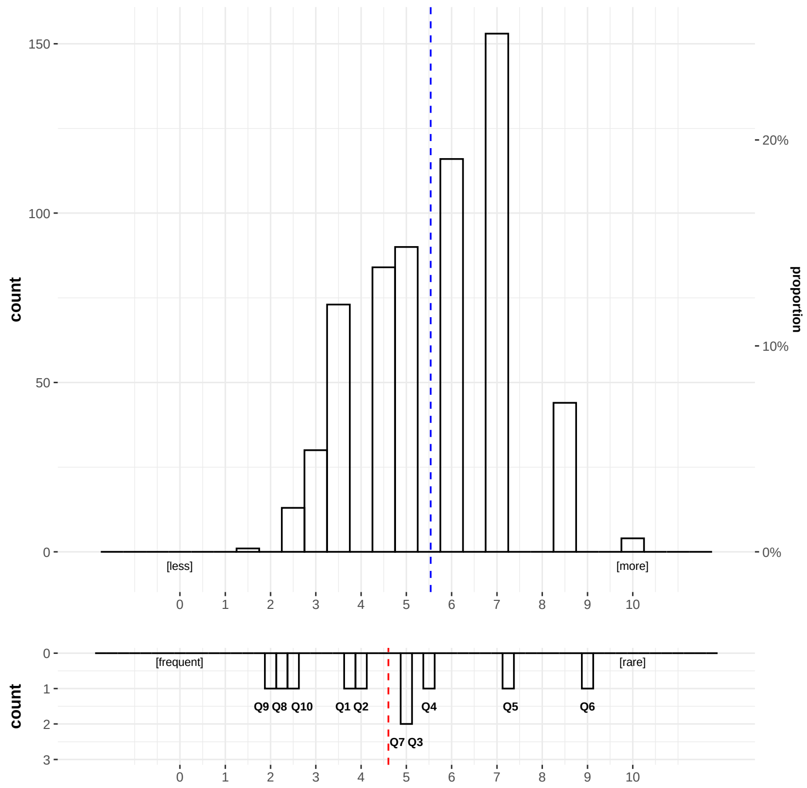


**Figure 1a Wright Map of Rater One scores**

**Figure 1b Wright Map of Rater Two scores**

**Rasch model was used to compare the two independent sets of rater scores. The distribution of scores was modelled, both including and removing outliers. Removing the outliers improved the fit, but did not change the conclusions. The distributions confirms that the value added to each criteria by each rater is the same inferring consistency.**
